# Effectiveness of amoxicillin and amoxicillin-clavulanate for the treatment of community-acquired pneumonia in adults and children: systematic review and meta-analysis

**DOI:** 10.1101/2025.09.05.25335164

**Authors:** Lilia Potter-Schwartz, Myo Maung Maung Swe, Michael Sharland, Julia Anna Bielicki, Ben S Cooper, Cherry Lim

## Abstract

**Introduction:** Community-acquired pneumonia (CAP) is a key contributor to the global burden of morbidity and mortality, and it drives large volumes of antibiotic use. Despite widespread recommendation to use amoxicillin for treatment, there are concerns of its effectiveness as empirical antibiotics for CAP. The aim of this study is to evaluate existing evidence on the effectiveness of amoxicillin and amoxicillin-clavulanate for CAP in children and adults.

**Methods:** We systematically searched PubMed, Cochrane Library, Web of Science, and Ovid-MEDLINER with no language restrictions for randomized controlled trials published until July 16, 2024 comparing the effectiveness of amoxicillin or amoxicillin-clavulanate versus other antibiotics or placebo with a primary outcome of clinical resolution or clinical failure. We used a random effects and a fixed effects logistic regression model to estimate the pooled treatment effect size. We performed a network meta-analysis for the indirect comparison between amoxicillin and amoxicillin-clavulanate.

**Results:** We extracted data from 44 studies including 45,400 patients. We found no strong evidence of a differential effect on clinical resolution when comparing amoxicillin to other antibiotics (n=15 trials, pooled odds ratio [OR]: 0.88; 95% confidence interval [CI]: 0.56-1.38) or amoxicillin-clavulanate to other antibiotics (n=17 trials, OR: 0.89 [95%: 0.76-1.04]).

Similarly, no evidence on difference in clinical failure between amoxicillin and other antibiotics (n=8 trials, OR: 1.34 [95%: 0.31, 5.81]) was observed. Sixty-three percent and 23% of amoxicillin and amoxicillin-clavulanate studies, respectively, had low-risk of bias. We found no strong evidence of a differential effect on clinical resolution between adults treated with amoxicillin and amoxicillin-clavulanate (OR: 1.06 [95%: 0.59-1.89]).

**Conclusion:** Our study found no strong evidence of any difference in clinical outcomes with amoxicillin compared to amoxicillin-clavulanate or other antibiotics for CAP treatment.

However, there are large uncertainties around the pooled estimates.

**Key Messages:** *What is already known on this topic:*

1. Community-acquired pneumonia (CAP) is a major cause of hospitalizations and of deaths globally. It is one of the most common causes of childhood hospitalizations in developed countries and the leading cause of death among children in developing countries
2. The World Health Organization (WHO) recommends amoxicillin and amoxicillin-clavulanate as the first and second choice, respectively, of empirical antibiotic treatment for mild to moderate bacterial CAP due to their narrow spectrum of activity and affordability.

*What this study adds:*

- While clinical trials have been performed to evaluate the effectiveness and efficacy of both antibiotics across different settings, studies directly comparing amoxicillin and amoxicillin-clavulanate are lacking. This study fills the gap through a systematic review to evaluate evidence on effectiveness of amoxicillin and amoxicillin-clavulanate versus other treatments for CAP and an indirect comparison between the two antibiotics.
- No strong evidence of a differential treatment effect when comparing amoxicillin to other antibiotics or amoxicillin-clavulanate to other antibiotics, or of difference in treatment effect between adults treated with amoxicillin and amoxicillin-clavulanate.
- How this study might affect research, practice or policy
- This work provides a strong rationale for the WHO recommendation of using amoxicillin as a first-choice empirical antibiotic for CAP in both adults and children.

## Introduction

Lower respiratory infections were the fourth highest contributor to global disability-adjusted life-years (DALYs) in 2019 [1]. Specifically, pneumonia causes a high global burden of death and morbidity, disproportionately affecting children less than 5 years old [2]. In 2021, there were an estimated 0.5 million (95% UI 0.4-0.6) pneumonia deaths in children under five. Community-acquired pneumonia (CAP) is one of the most common causes of childhood hospitalizations in developed countries and the leading cause of death among children in developing countries [2– 4]. The empirical antibiotic treatment guidelines of CAP vary by country and can be based on severity scores and local epidemiology [5–7].

Amoxicillin is recommended by the World Health Organization (WHO) as the first-choice antibiotic for adults and children in 10 of the 12 most common primary care infections for all WHO regions, making it a critical ‘Access’ antibiotic. In particular, it is recommended as the first choice for mild/moderate bacterial CAP in adults, with amoxicillin-clavulanate (amoxicillin + clavulanate) as the second choice antibiotic [8,9]. Amoxicillin is also the WHO recommended first choice treatment for all children with non-severe pneumonia [8,9]. Similarly, for the WHO AWaRe book, amoxicillin is the first choice for bacterial CAP in adults and children, while amoxicillin-clavulanate is a second choice recommendation [8,9]. Improving the evidence base for the use of amoxicillin empirically for CAP could help support its key role in achieving the global United Nation General Assembly antimicrobial resistance (UNGA AMU) target of 70% of overall human antibiotic consumption from the ‘Access’ group by 2030.

While clinical trials have examined amoxicillin across different settings, studies directly comparing amoxicillin and amoxicillin-clavulanate are lacking. We performed a systematic review on trials of amoxicillin or amoxicillin-clavulanate and evaluated the effectiveness in empirical treatments for CAP between the two regimens and other antibiotics using a network meta-analysis.

## Methods

### Search Strategy

Our search strategy of randomized trials across four databases used keywords pertinent to the evaluation of amoxicillin and amoxicillin-clavulanate in treatment of community-acquired pneumonia. We searched PubMed, Cochrane Library, Web of Science, and Ovid-MEDLINER databases using the following search term: ((amoxicillin[Title/Abstract] OR amoxicillin-clavulanate[Title/Abstract] OR amoxicillin/clavulanic acid[Title/Abstract] OR amoxycillin[Title/Abstract] OR hydroxyampicillin[Title/Abstract] OR amoxil[Title/Abstract] OR clamoxyl[Title/Abstract] OR trimox[Title/Abstract] OR wymox[Title/Abstract] OR polymox[Title/Abstract] OR larotid[Title/Abstract] OR augmentin[Title/Abstract]) AND (pneumonia[Title]) NOT (review[Title])).

Each source was last searched on July 16, 2024 and reference lists of the included articles were screened, inclusive of articles published in any language. Our review followed a prespecified protocol and was registered with the International Prospective Register of Systematic Reviews (PROSPERO registration number: CRD42024568554. We followed the Preferred Reporting Items for Systematic Reviews and Meta-analysis (PRISMA) guidelines [10].

### Eligibility Criteria

Studies were eligible if i) they were randomized controlled trials (RCTs), ii) the objectives were to compare amoxicillin or amoxicillin-clavulanate to another antibiotic or placebo for either outpatients or inpatients diagnosed with CAP, iii) published prior to June 2024. Observational studies, qualitative studies, case reports, and review were excluded. Additionally, studies were excluded if patients with other infections besides CAP were randomized, multiple antibiotics were used in the amoxicillin arm (i.e. combined treatment of amoxicillin with other antibiotics), or the primary outcome was neither clinical resolution nor clinical failure.

### Study Selection

We first removed duplicated studies, and then screened the titles and abstracts. Two independent reviewers (LPS, MMMS) performed the screening to select articles for a full review. Any disagreements between the reviewers were resolved by a third reviewer (CL). A full article review was then conducted for each selected study. We recorded the reasons for excluding studies (Figure 1, https://github.com/liliapsnh/cap_amoxicillin.git).

**Figure 1.**
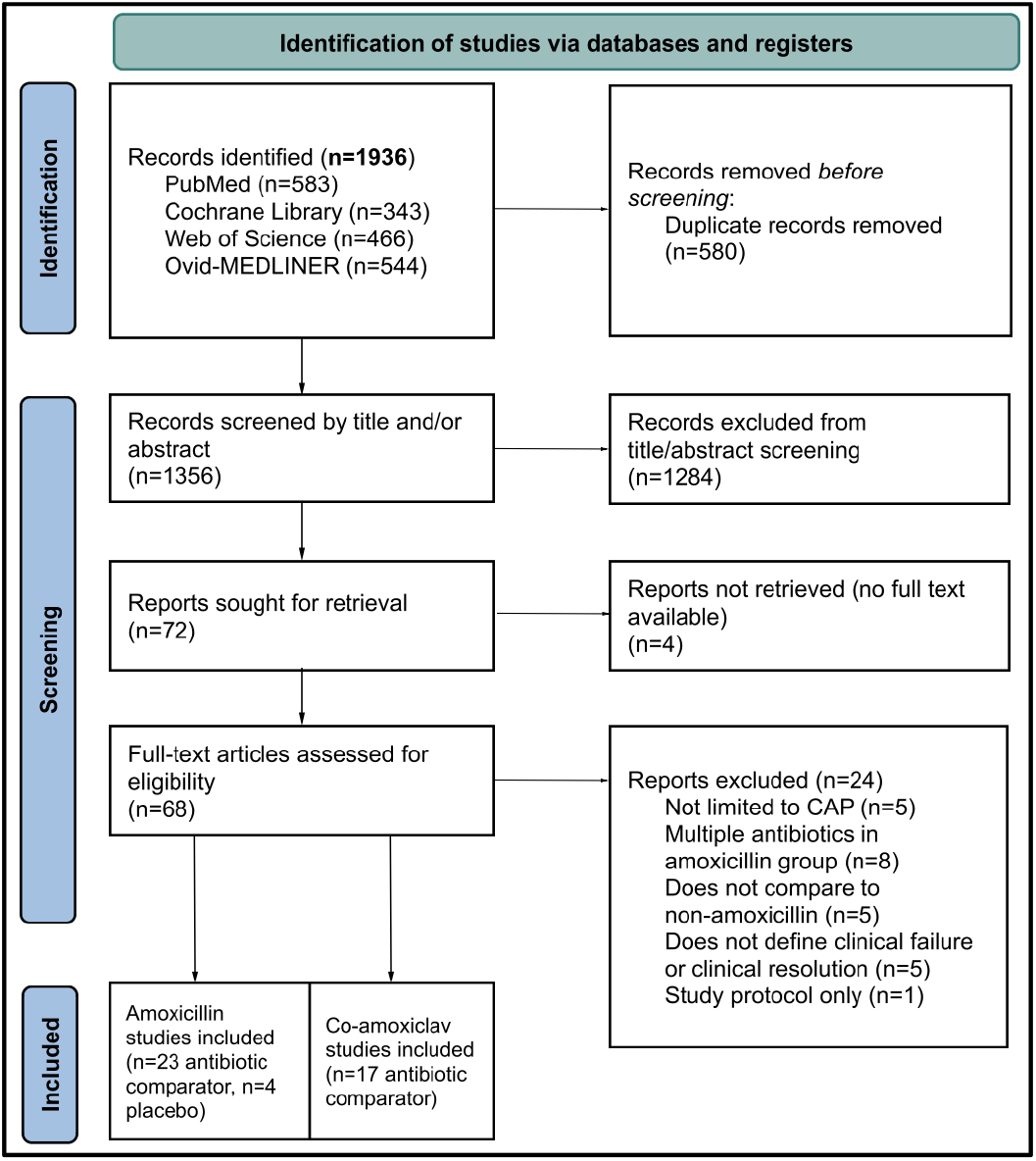
PRISMA 2020 flow diagram for systematic reviews.

### Data Extraction

We developed a data extraction spreadsheet with the intended variables. After piloting data extraction for five randomly selected articles meeting inclusion criteria, the spreadsheet was finalised using feedback from four reviewers (LPS, MMMS, BC, CL). The list of extracted variables included title, first author, journal, PMID, DOI, publication year, study type, setting, study blinding, CAP criteria, patient population, geographical region, sample size, primary and secondary clinical outcomes, drug adherence, patients lost to follow-up, adverse events, and type of analysis (intention-to-treat or per-protocol). The antibiotic name, dose, and route of administration for both the intervention and comparator treatments were also extracted. The primary outcomes extracted from the included trials were clinical resolution and clinical failure, with a secondary outcome of microbiological success (confirmed or presumed bacteriological eradication).

### Risk of Bias Assessment

The revised Cochrane risk of bias tool for randomized trials (RoB2) was used to assess and categorise studies into low, some concerns, or high risk of bias. The five domains of the RoB2 tool assessed the randomisation process, deviations from intended interventions, missing outcome data, measurement of the outcome, and selection of the reported result. Two reviewers (LPS, MMMS) independently evaluated the studies using the RoB2 tool, with a third reviewer (CL) resolving any conflicts that arose.

### Statistical Analysis

We used the meta and netmeta packages in R (version 4.4.0) to perform the meta-analyses [11]. The meta package was used to calculate pooled odds ratios and generate forest plots. The netmeta package was used for the network plots and network meta-analysis. We evaluated the effect of amoxicillin against alternative antibiotics on outcomes using a random effects logistic regression model to include between-study variability into the estimation of the overall treatment effects. We then evaluated the effect of amoxicillin against placebo on outcomes using fixed-effects logistic regression models. The effect sizes were reported as a pooled odds ratio (OR).

Separate models were fitted on data from studies that included amoxicillin-clavulanate in one of the treatment arms. Two primary outcomes, which are clinical resolution and treatment failure, were analysed in separate models. Secondary outcome analysis for bacterial eradication was also performed. We performed subgroup analyses by age group (<5 and >5 years), disease severity, antibiotic class, and dose, following the prespecified protocol.

Finally, we performed network meta-analysis to compare amoxicillin versus amoxicillin-clavulanate, assuming random effects. To demonstrate the utility of network meta-analysis for this analysis, we simulated 50 studies of 1:1 randomized controlled trials for six different treatments (x = 1, …, 6) of disease Y. A network meta-analysis was performed on the 50 studies to compare the effectiveness of treatments x = 1 and x = 2. A separate network meta-analysis was performed on the subcohort of those 50 studies that included at least one of the two treatments (k=1 and/or k=2) of interest. The code for the simulation studies and the results from the two network meta-analyses on the simulated data are available in https://github.com/Cherrylim128/AmoxForCap.

We also evaluated the effect of amoxicillin against other antibiotic treatment on outcomes using fixed-effects logistic regression models as a sensitivity analyses, assuming there was a true underlying effect of amoxicillin on the outcome across the studies included in the meta-analysis.

Heterogeneity of the studies was evaluated using the *I^2^*statistic. The statistical heterogeneity expressed with the *I^2^* statistic conveys the percent of variability in the effect estimates that is attributed to heterogeneity instead of sampling error [12].

## Results

### Study Characteristics

We identified 1,936 records across four databases (PubMed, Cochrane Library, Web of Science, and Ovid-MEDLINER) and after removing duplicates we screened 1,356 abstracts, of which 27 studies on amoxicillin and 17 studies on amoxicillin-clavulanate met the eligibility criteria and were included in the final review and meta-analysis (Figure 1, Supplementary Figure 1). Of the 27 randomized controlled trials comparing amoxicillin to other antibiotics or placebo, 15 and 12 reported primary outcomes for clinical resolution and clinical failure, respectively, for patients with CAP. The 17 studies comparing amoxicillin-clavulanate to other antibiotics reported a primary outcome of clinical resolution.

The amoxicillin and amoxicillin-clavulanate randomized controlled trials included in the review represented 39,968 and 5,432 patients, respectively, ranging from 32 to 15,662 patients per trial (Supplementary Table 1). There were six amoxicillin-clavulanate three-arm studies that compared either two different dosages of the same antibiotic (levofloxacin, cefditoren) or two other different antibiotics to amoxicillin-clavulanate within the same study. Four of the amoxicillin studies were placebo-controlled, and all four trials were on patients less than five years of age with non-severe diagnoses. The dose and duration of amoxicillin and amoxicillin-clavulanate treatment varied by study, as well as the clinical severity, country, and study population (https://github.com/liliapsnh/cap_amoxicillin.git). The duration of amoxicillin ranged from 2-14 days and the dosage ranged from 1-4 grams/day for adult patients.

### Comparator Antibiotics

The trials included in this systematic review compared amoxicillin to 15 different antibiotics and amoxicillin-clavulanate to 13 different antibiotics. By antibiotic class, the trials included treatment arms that compared amoxicillin to beta-lactams (n=8), macrolides (n=3), quinolones (n=6), and sulfonamides (n=6); and amoxicillin-clavulanate to beta-lactams (n=10), macrolides (n=7), and quinolones (n=7). The six studies with sulfonamides all compared oral amoxicillin to oral co-trimoxazole in study populations of children less than five years of age with non-severe CAP. After co-trimoxazole, the most frequent pairs of comparator antibiotics included amoxicillin-clavulanate to ceftriaxone (n=4), amoxicillin-clavulanate to cefuroxime (n=3), amoxicillin-clavulanate to erythromycin (n=3), and amoxicillin to benzylpenicillin (n=3).

### Primary and Secondary Outcomes

The definition of clinical resolution used in the trials was resolution of symptoms (29 studies) or no requirement for further antibiotic therapy (three studies). Five amoxicillin and three amoxicillin-clavulanate studies included confirmation from chest x-ray results at test of cure visits. The clinical resolution outcome was examined at various time points ranging from day 5 of treatment to 25 days post therapy. The random effects model did not find strong evidence of differences in the odds of clinical resolution between amoxicillin treatment compared to other antibiotic treatment (n=15; pooled odds ratio [OR]: 0.88; 95% confidence interval [CI]: 0.56-1.38; I^2^=71%) (Figure 2) and amoxicillin-clavulanate compared to other antibiotics (n=17; pooled OR: 0.89, 95% CI: 0.76-1.04; I^2^=0%) (Figure 3).

**Figure 2.**
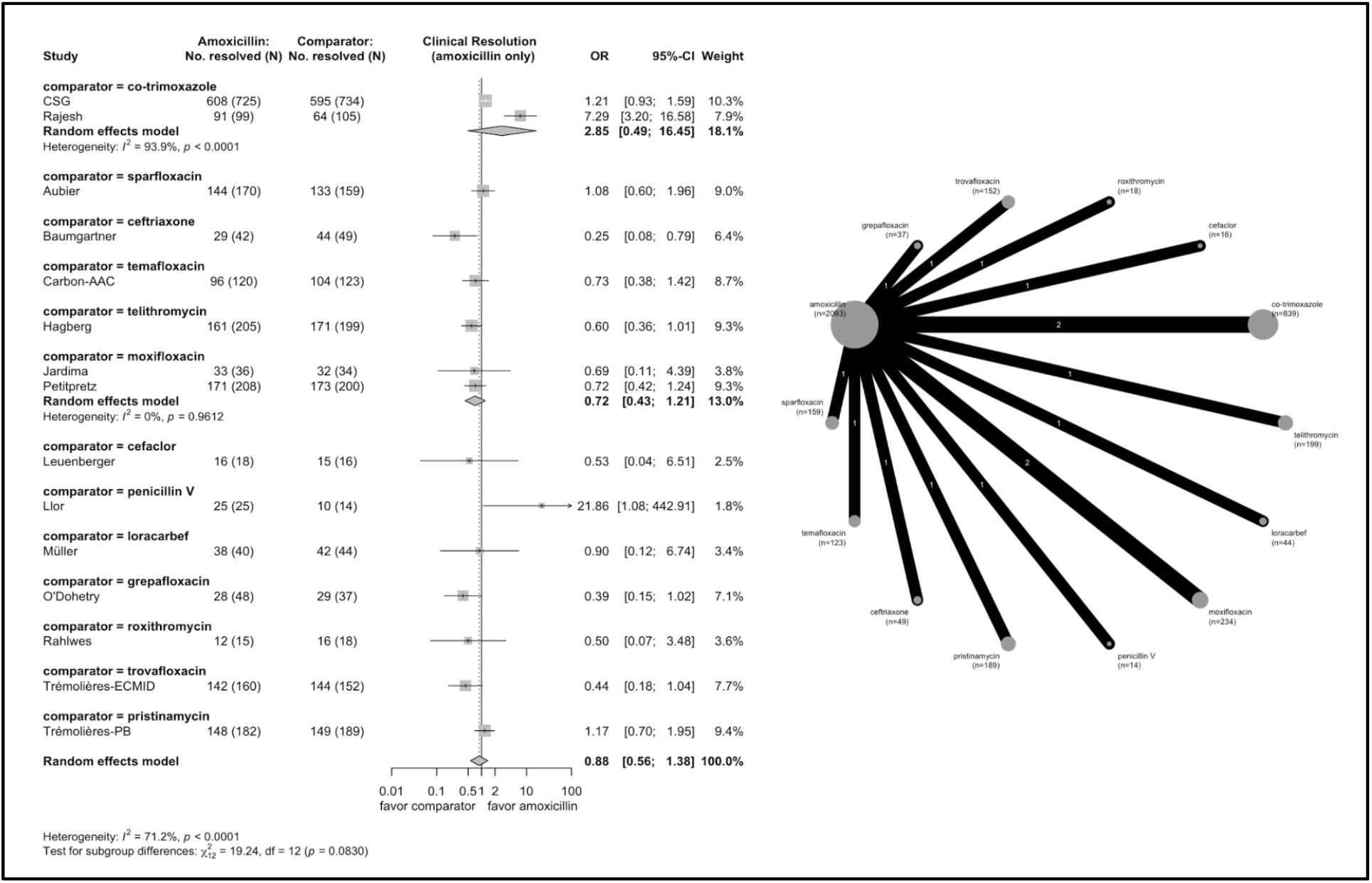
Forest plot and network graph for the 15 studies with a primary outcome of clinical resolution comparing amoxicillin to another antibiotic treatment. Two studies (CSG, Rajesh) consider a patient population less than five years of age. Larger area of the circle in the network diagram reflects a higher number of papers using the antibiotic in one of the treatment arms.

**Figure 3.**
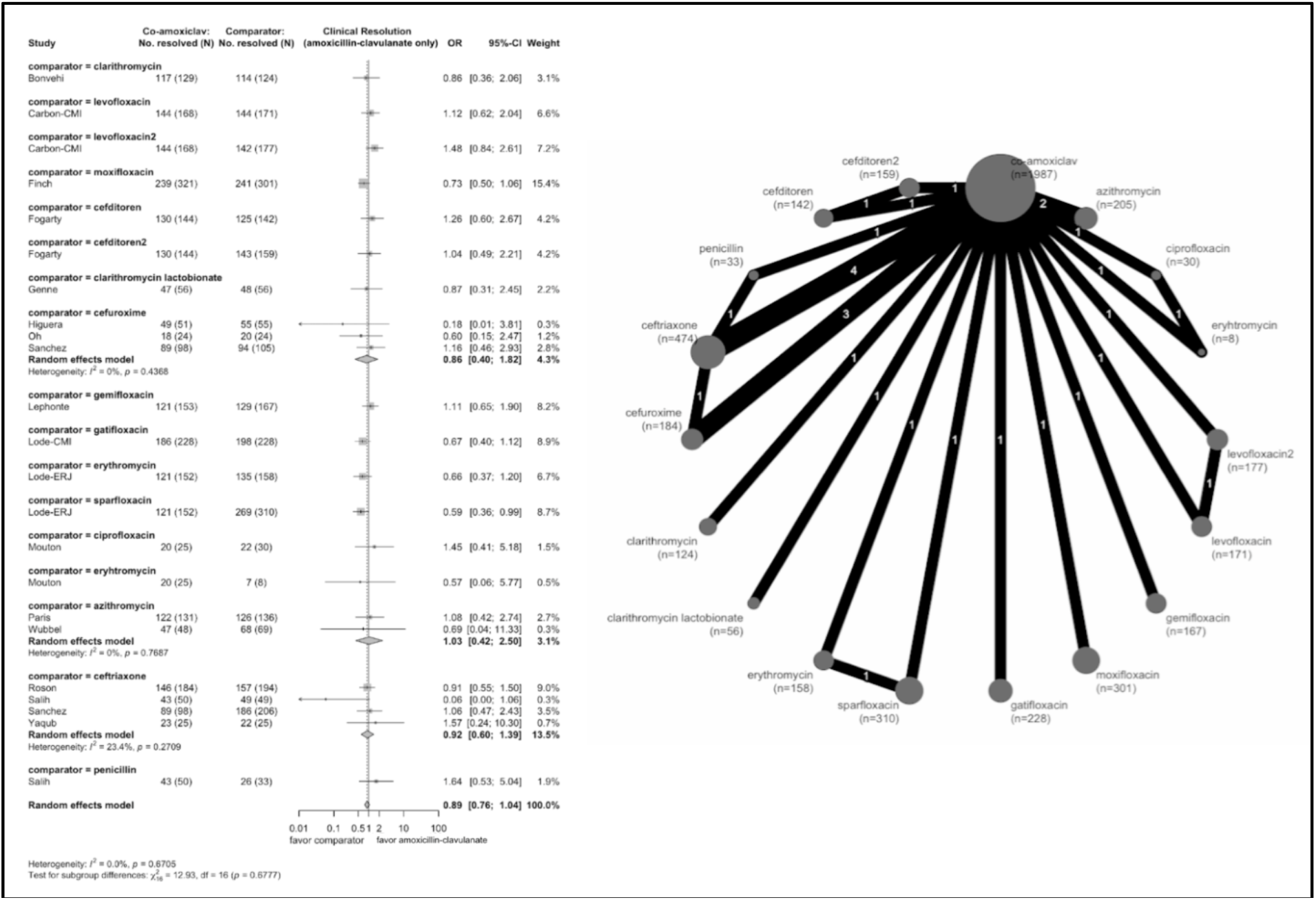
Forest plot and network graph for the 17 studies with a primary outcome of clinical resolution comparing amoxicillin-clavulanate to another antibiotic treatment. Two studies (Salih, Wubbel) considers a patient population less than five years of age.

Twelve of the 27 studies with an amoxicillin treatment arm analysed clinical failure as the primary outcome. In 10 studies the definition of clinical failure was persistence of symptoms while in two studies it was the requirement to change antibiotic regimen. The clinical failure outcome was examined at various time points ranging from 48 hours to 14 days after starting the treatment regimen. There were eight trials comparing clinical failure in amoxicillin to another antibiotic treatment (non-placebo trial), with large heterogeneity. The reported OR from those trials ranged from 0.66 [95% CI: 0.5, 0.86] to 2.52 [95% CI: 2.21, 2.87] (Figure 4).

**Figure 4.**
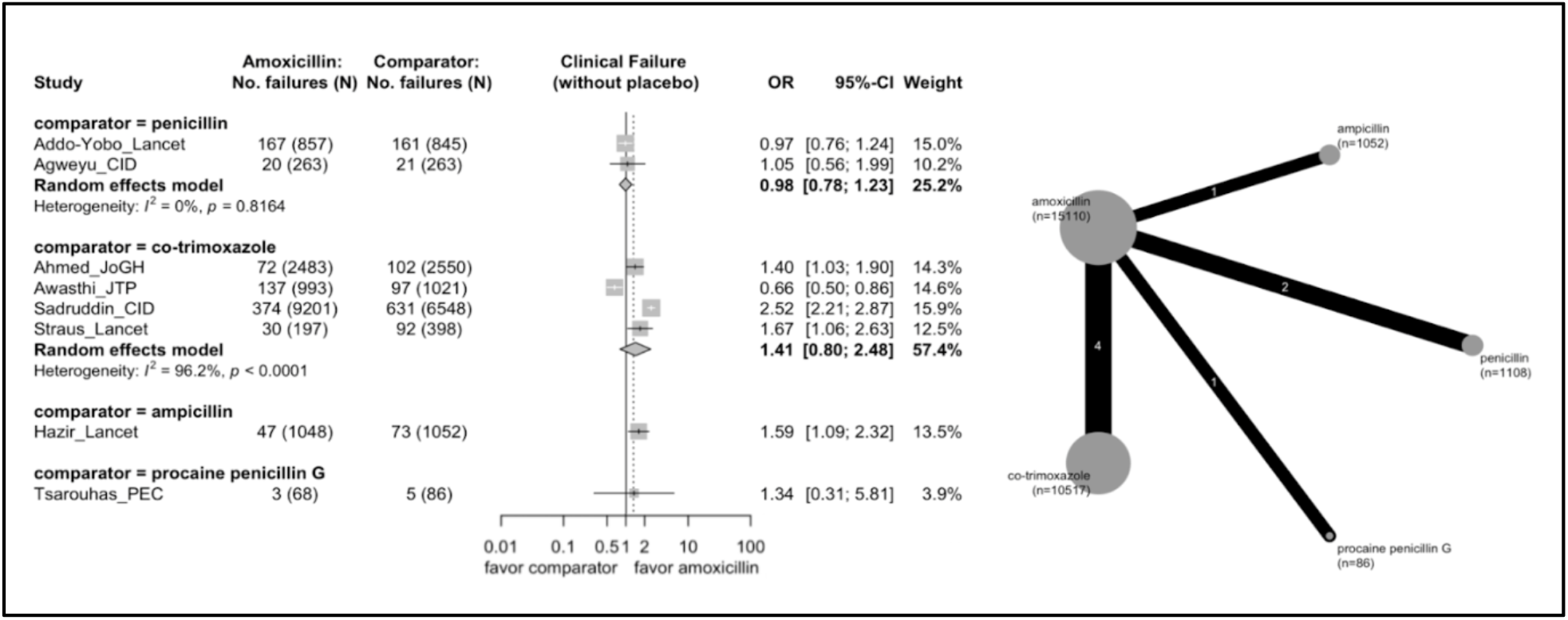
Forest plot and network graph for the 8 studies with a primary outcome of clinical failure comparing amoxicillin to another antibiotic (non-placebo). One study (Tsarouhas) considers a patient population of greater than five years of age.

There were 28 studies evaluating adult patients with an outcome of clinical resolution and network meta-analysis on those studies found no strong evidence of differential effect on outcome between adults treated with amoxicillin versus those treated with amoxicillin-clavulanate (pooled OR: 1.06 [95% CI: 0.59, 1.89]) (Supplementary Figure 2).

Thirteen studies reported bacteriological success eradication as a secondary outcome for patients with available microbiological samples. The pooled odds ratio for the studies comparing amoxicillin to alternative antibiotics was 0.53 [95% CI: 0.32, 0.87] (Supplementary Figure 3) and for studies comparing amoxicillin-clavulanate to alternative antibiotics it was 0.80 [95% CI: 0.53, 1.22] (Supplementary Figure 4).

The fixed effects models, under the assumption that there was a true underlying effect across all studies, yielded narrower confidence intervals (Supplementary Figures 5-6). A clinical benefit of amoxicillin compared to placebo in four studies was observed with an estimated pooled odds ratio of 1.37 [95% CI: 1.16, 1.62] from the fixed effects model (Figure 5). All of the placebo studies had a duration of oral amoxicillin treatment for three days with dosage between 31 mg/kg/day and 1000mg/day [18,22,25,27].

**Figure 5.**
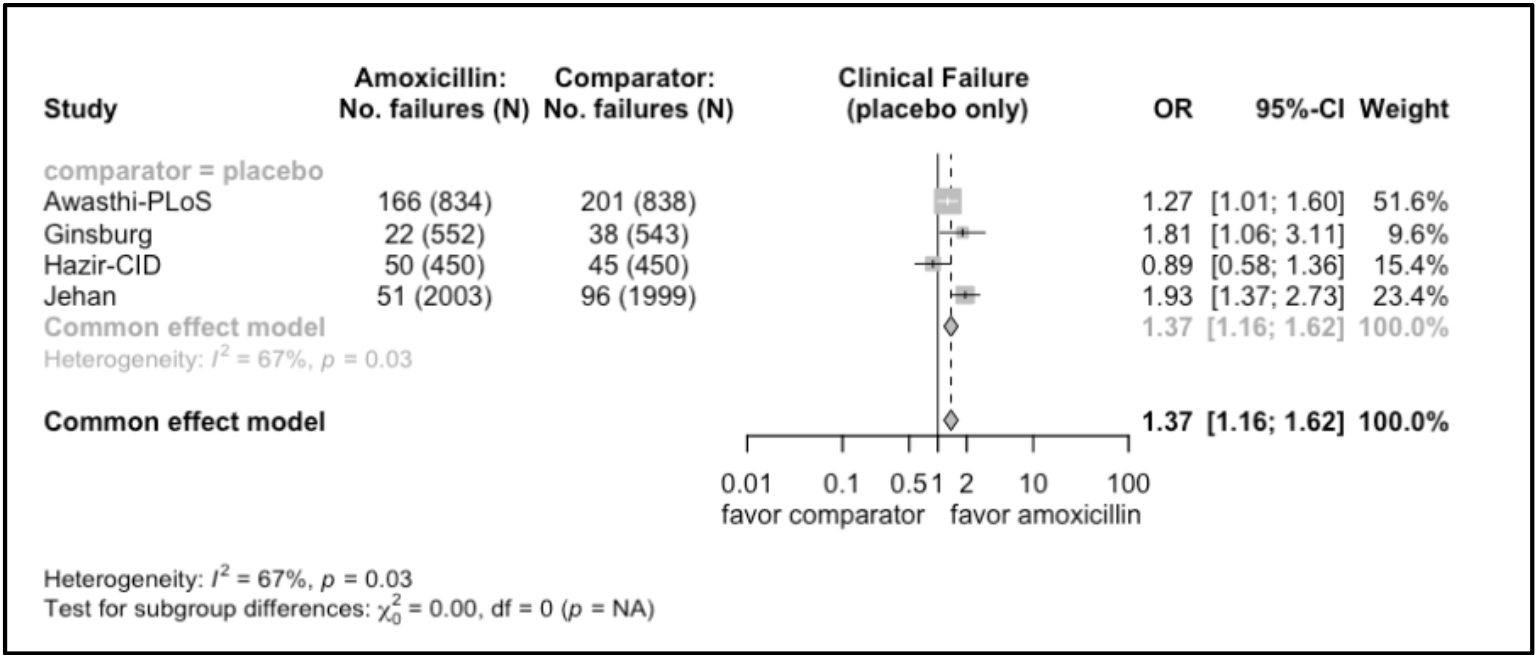
Forest plot for fixed effects model of clinical failure and placebo-comparator only.

### Subgroup Analyses

We stratified the included studies by clinical outcome, age group, antibiotic class, and amoxicillin dose. Thirteen (48%) of the amoxicillin and two (12%) of the amoxicillin-clavulanate studies assessed patient populations less than five years of age. We observed similar patterns across subgroup analyses by antibiotic class (Supplementary Figures 7-9) and amoxicillin or amoxicillin-clavulanate dose (Supplementary Figures 10-15).

### Risk of Bias Assessment

The distribution of overall risk of bias scores among the amoxicillin studies were 63% (n=17) low risk, 26% (n=7) some concerns, and 11% (n=3) high risk (Supplementary Figure 16). The distribution of scores for amoxicillin-clavulanate studies was 29% (n=5) low risk, 47% (n=8) some concerns, and 24% (n=4) high risk (Supplementary Figure 16). The most common reasons for concerns of bias included limited information about the randomization process, a high proportion of patients (>10%) lost to follow-up, and failure to report results on an intention to treat basis.

## Discussion

Our systematic review and meta-analysis suggested that of the 44 trials identified, there is no strong evidence of a differential effectiveness of either amoxicillin or amoxicillin-clavulanate compared to other antibiotic treatments. The four identified placebo trials for amoxicillin, performed in India, Malawi, and Pakistan (23, 27, 30, 32) highlighted the effectiveness of amoxicillin among children with non-severe CAP in low and middle-income countries and point to the importance of ensuring sustainable access to amoxicillin.

The wide confidence interval around the treatment effect for amoxicillin compared to other antibiotics reflected large variations across the trials. The point estimates from the main analyses and sub-group analyses ranged from 0.75 to 1.41 (Supplementary Figure 9, Supplementary Figure 13) indicating that reported differences in effectiveness between amoxicillin and other antibiotics for CAP were small. A population-based emulated target trial that compared 25,332 cases of paediatric pneumonia in Sweden showed lower risk of treatment failure in the amoxicillin treated group compared to the penicillin V treated group [57].

We found no strong evidence of difference in effectiveness between amoxicillin-clavulanate and amoxicillin using a network meta-analysis. This finding is consistent with a retrospective observational study of electronic health records, which found no evidence of a difference in mortality between the two treatments for CAP [58]. Similarly, a multi-centre randomized trial conducted in seven hospitals in the Netherlands showed narrow-spectrum treatment for non-ICU CAP was non-inferior to broad-spectrum treatment [59]. There is increasing evidence of the importance of using beta-lactam monotherapies, such as amoxicillin, in the treatment of CAP to reduce the use of macrolides and fluoroquinolones. This is due to concerns about potentially increased selection for resistance with Watch or Reserve antibiotics, which was suggested by their stronger association with multidrug-resistant organisms compared to Access antibiotics. [60].

Our subgroup analysis focusing on trials using high-dose amoxicillin (≥80 mg/kg/day in children or ≥3000 mg/day in adults) found no evidence of a differential effect compared to antibiotics.

This finding is consistent with the observations from the recent clinical trials that focused on effective duration and dose of amoxicillin for community-acquired paediatric pneumonia, suggesting similar treatment outcomes between low and high dose amoxicillin [61–64].

Limitations of our study included the insufficient number of trials to comprehensively adjust for differences in amoxicillin doses and treatment duration in the main analysis. There was also inconsistent reporting on microbiological cure rates in the included trials and heterogeneity in the type of comparator antibiotics used across the trials identified, which could explain the observed variations in the treatment effect. Our network meta-analysis was based on trials focusing on amoxicillin or amoxicillin-clavulanate only. There may be other relevant trials that could contribute to the indirect evidence for the comparison between amoxicillin and amoxicillin-clavulanate than those included in this analysis. Future studies would be needed to strengthen the evidence for the recommendations for the choice between those two antibiotics for empiric treatment in CAP.

## Conclusion

In conclusion, our findings provide a strong rationale for the WHO AWaRe book recommendation of using amoxicillin as a first-choice antibiotic for the treatment of CAP in adults and children. The findings from our study also support the widespread recommendation of using amoxicillin as the empirical antibiotic treatment for outpatients without comorbidities diagnosed with CAP across different local and national antimicrobial treatment guidelines.

## Data Availability

All data produced in the present work are contained in the manuscript or available online.

## References

1. GBD 2019 Diseases and Injuries Collaborators. Global burden of 369 diseases and injuries in 204 countries and territories, 1990-2019: a systematic analysis for the Global Burden of Disease Study 2019. Lancet. 2020;396:1204–22.

2. UNICEF. A child dies of pneumonia every 43 seconds. 2024. https://data.unicef.org/topic/child-health/pneumonia/#:~:text=Pneumonia (accessed 29 August 2024)

3. Meyer Sauteur PM. Childhood community-acquired pneumonia. Eur J Pediatr. 2024;183:1129–36.

4. Number of deaths from pneumonia in children under five. Our World in Data. 2024. https://ourworldindata.org/grapher/number-of-deaths-from-pneumonia-in-children-under-5?tab=table (accessed 15 January 2025)

5. Eccles S, Pincus C, Higgins B, et al. Diagnosis and management of community and hospital acquired pneumonia in adults: summary of NICE guidance. BMJ. 2014;349:g6722.

6. Höffken G, Lorenz J, Kern W, et al. Guidelines of the Paul-Ehrlich-society of chemotherapy, the German respiratory diseases society, the German infectious diseases society and of the competence network CAPNETZ for the management of lower respiratory tract infections and community-acquired pneumonia. Pneumologie. 2010;64:149–54.

7. Jain S, Self WH, Wunderink RG, et al. Community-acquired pneumonia requiring hospitalization among U.S. adults. N Engl J Med. 2015;373:415–27.

8. Sharland M, Cook A, Pouwels KB, et al. Universal access to key essential antibiotics— Recent amoxicillin global shortages mask a wider policy failure. CMI Communications. 2024;1:105035.

9. The WHO AWaRe (Access, Watch, Reserve) antibiotic book. 2022. https://www.who.int/publications/i/item/9789240062382 (accessed 13 August 2024)

10. Page MJ, McKenzie JE, Bossuyt PM, et al. The PRISMA 2020 statement: an updated guideline for reporting systematic reviews. BMJ. 2021;372:n71.

11. _R: A Language and Environment for Statistical Computing_. R Foundation for Statistical Computing. 2024.

12. Chapter 10: Analysing data and undertaking meta-analyses. https://training.cochrane.org/handbook/current/chapter-10 (accessed 3 November 2024)

13. Catchup Study Group. Clinical efficacy of co-trimoxazole versus amoxicillin twice daily for treatment of pneumonia: a randomized controlled clinical trial in Pakistan. Arch Dis Child. 2002;86:113–8.

14. Addo-Yobo E, Chisaka N, Hassan M, et al. Oral amoxicillin versus injectable penicillin for severe pneumonia in children aged 3 to 59 months: a randomized multicentre equivalency study. Lancet. 2004;364:1141–8.

15. Agweyu A, Gathara D, Oliwa J, et al. Oral Amoxicillin Versus Benzyl Penicillin for Severe Pneumonia Among Kenyan Children: A Pragmatic Randomized Controlled Noninferiority Trial. Clin Infect Dis. 2015;60:1216.

16. Ahmed S, Ariff S, Muhammed S, et al. Community case management of fast-breathing pneumonia with 3 days oral amoxicillin vs 5 days cotrimoxazole in children 2-59 months of age in rural Pakistan: A cluster randomized trial. J Glob Health. 2022;12:04097.

17. Aubier M, Verster R, Regamey C, et al. Once-daily sparfloxacin versus high-dosage amoxicillin in the treatment of community-acquired, suspected pneumococcal pneumonia in adults. Sparfloxacin European Study Group. Clin Infect Dis. 1998;26:1312–20.

18. Awasthi S, Agarwal G, Kabra SK, et al. Does 3-day course of oral amoxycillin benefit children of non-severe pneumonia with wheeze: a multicentric randomized controlled trial. PLoS One. 2008;3:e1991.

19. Awasthi S, Agarwal G, Singh JV, et al. Effectiveness of 3-day amoxycillin vs. 5-day co-trimoxazole in the treatment of non-severe pneumonia in children aged 2-59 months of age: a multi-centric open labeled trial. J Trop Pediatr. 2008;54:382–9.

20. Baumgartner JD, Glauser MP. Tolerance study of ceftriaxone compared with amoxicillin in patients with pneumonia. Am J Med. 1984;77:54–8.

21. Carbon C, Léophonte P, Petitpretz P, et al. Efficacy and safety of temafloxacin versus those of amoxicillin in hospitalized adults with community-acquired pneumonia. Antimicrob Agents Chemother. 1992;36:833–9.

22. Ginsburg AS, Mvalo T, Nkwopara E, et al. Placebo vs Amoxicillin for Nonsevere Fast-Breathing Pneumonia in Malawian Children Aged 2 to 59 Months: A Double-blind, Randomized Clinical Noninferiority Trial. JAMA Pediatr. 2019;173:21–8.

23. Hagberg L, Torres A, van Rensburg D, et al. Efficacy and tolerability of once-daily telithromycin compared with high-dose amoxicillin for treatment of community-acquired pneumonia. Infection. 2002;30:378–86.

24. Hazir T, Fox LM, Nisar YB, et al. Ambulatory short-course high-dose oral amoxicillin for treatment of severe pneumonia in children: a randomized equivalency trial. Lancet. 2008;371:49–56.

25. Hazir T, Nisar YB, Abbasi S, et al. Comparison of oral amoxicillin with placebo for the treatment of world health organization-defined nonsevere pneumonia in children aged 2-59 months: a multicenter, double-blind, randomized, placebo-controlled trial in Pakistan. Clin Infect Dis. 2011;52:293–300.

26. Jardim JR, Rico G, de la Roza C, et al. [A comparison of moxifloxacin and amoxicillin in the treatment of community-acquired pneumonia in Latin America: results of a multicenter clinical trial]. Arch Bronconeumol. 2003;39:387–93.

27. Jehan Fyezah, Nisar Imran, Kerai Salima, et al. Randomized Trial of Amoxicillin for Pneumonia in Pakistan. N Engl J Med. 2020;383:24–34.

28. Leuenberger P, Vrantchev S. Cefaclor versus amoxicillin in the treatment of bacterial pneumonia: a comparative double-blind study. Eur J Clin Microbiol. 1983;2:11–6.

29. Llor C, Pérez A, Carandell E, et al. Efficacy of high doses of penicillin versus amoxicillin in the treatment of uncomplicated community acquired pneumonia in adults. A non-inferiority controlled clinical trial. Aten Primaria. 2019;51:32–9.

30. Müller O, Wettich K. Comparison of loracarbef (LY163892) versus amoxicillin in the treatment of bronchopneumonia and lobar pneumonia. Infection. 1992;20:176–82.

31. O’Doherty B, Dutchman DA, Pettit R, et al. Randomized, double-blind, comparative study of grepafloxacin and amoxycillin in the treatment of patients with community-acquired pneumonia. J Antimicrob Chemother. 1997;40 Suppl A:73–81.

32. Petitpretz P, Arvis P, Marel M, et al. Oral moxifloxacin vs high-dosage amoxicillin in the treatment of mild-to-moderate, community-acquired, suspected pneumococcal pneumonia in adults. Chest. 2001;119:185–95.

33. M Rahlwes, J Wagner, L Schuster, E Berntsson, G Ruckdeschel, W Ehret, M Priesnitz, H Lode. Prospective randomized comparison of roxithromycin and amoxycillin in non-hospital-acquired pneumonia. Br J Clin Pract. 1988;42:91.

34. Rajesh SM, Singhal V. Clinical Effectiveness of Co-trimoxazole vs. Amoxicillin in the Treatment of Non-Severe Pneumonia in Children in India: A Randomized Controlled Trial. Int J Prev Med. 2013;4:1162–8.

35. Sadruddin S, Khan IUH, Fox MP, et al. Comparison of 3 Days Amoxicillin Versus 5 Days Co-Trimoxazole for Treatment of Fast-breathing Pneumonia by Community Health Workers in Children Aged 2-59 Months in Pakistan: A Cluster-randomized Trial. Clin Infect Dis. 2019;69:397–404.

36. Straus WL, Qazi SA, Kundi Z, et al. Antimicrobial resistance and clinical effectiveness of co-trimoxazole versus amoxycillin for pneumonia among children in Pakistan: randomized controlled trial. Pakistan Co-trimoxazole Study Group. Lancet. 1998;352:270–4.

37. Trémolières F, de Kock F, Pluck N, et al. Trovafloxacin versus high-dose amoxicillin (1 g three times daily) in the treatment of community-acquired bacterial pneumonia. Eur J Clin Microbiol Infect Dis. 1998;17:447–53.

38. Trémolières F, Mayaud C, Mouton Y, et al. Efficacy and safety of pristinamycin vs amoxicillin in community acquired pneumonia in adults. Pathol Biol. 2005;53:503–10.

39. Tsarouhas N, Shaw KN, Hodinka RL, et al. Effectiveness of intramuscular penicillin versus oral amoxicillin in the early treatment of outpatient pediatric pneumonia. Pediatr Emerg Care. 1998;14:338–41.

40. Bonvehi P, Weber K, Busman T, et al. Comparison of Clarithromycin and Amoxicillin/Clavulanic Acid for Community-Acquired Pneumonia in an Era of Drug-Resistant Streptococcus pneumoniae. Clin Drug Investig. 2003;23:491–501.

41. Carbon C, Ariza H, Rabie WJ, et al. Comparative study of levofloxacin and amoxycillin/clavulanic acid in adults with mild-to-moderate community-acquired pneumonia. Clin Microbiol Infect. 1999;5:724–32.

42. Finch R, Schürmann D, Collins O, et al. Randomized controlled trial of sequential intravenous (i.v.) and oral moxifloxacin compared with sequential i.v. and oral co-amoxiclav with or without clarithromycin in patients with community-acquired pneumonia requiring initial parenteral treatment. Antimicrob Agents Chemother. 2002;46:1746–54.

43. Fogarty CM, Cyganowski M, Palo WA, et al. A comparison of cefditoren pivoxil and amoxicillin/clavulanate in the treatment of community-acquired pneumonia: a multicenter, prospective, randomized, investigator-blinded, parallel-group study. Clin Ther. 2002;24:1854–70.

44. Genné D, Siegrist HH, Humair L, et al. Clarithromycin versus amoxicillin-clavulanic acid in the treatment of community-acquired pneumonia. Eur J Clin Microbiol Infect Dis. 1997;16:783–8.

45. Higuera F, Hidalgo H, Feris J, et al. Comparison of oral cefuroxime axetil and oral amoxycillin/clavulanate in the treatment of community-acquired pneumonia. J Antimicrob Chemother. 1996;37:555–64.

46. Léophonte P, File T, Feldman C. Gemifloxacin once daily for 7 days compared to amoxicillin/clavulanic acid thrice daily for 10 days for the treatment of community-acquired pneumonia of suspected pneumococcal origin. Respir Med. 2004;98:708–20.

47. Lode H, Garau J, Grassi C, et al. Treatment of community-acquired pneumonia: a randomized comparison of sparfloxacin, amoxycillin-clavulanic acid and erythromycin. Eur Respir J. 1995;8:1999–2007.

48. Lode H, Magyar P, Muir JF, et al. Once-daily oral gatifloxacin vs three-times-daily co-amoxiclav in the treatment of patients with community-acquired pneumonia. Clin Microbiol Infect. 2004;10:512–20.

49. Mouton Y, Beuscart C, Leroy O, et al. Evaluation of ciprofloxacin versus amoxicillin + clavulanic acid or erythromycin for the empiric treatment of community-acquired pneumonia. Pathol Biol. 1991;39:34–7.

50. Oh HM, Ng AW, Lee SK. Cefuroxime compared to amoxicillin-clavulanic acid in the treatment of community-acquired pneumonia. Singapore Med J. 1996;37:255–7.

51. Paris R, Confalonieri M, Dal Negro R, et al. Efficacy and safety of azithromycin 1 g once daily for 3 days in the treatment of community-acquired pneumonia: an open-label randomized comparison with amoxicillin-clavulanate 875/125 mg twice daily for 7 days. J Chemother. 2008;20:77–86.

52. Rosón B, Carratalà J, Tubau F, et al. Usefulness of betalactam therapy for community-acquired pneumonia in the era of drug-resistant Streptococcus pneumoniae: a randomized study of amoxicillin-clavulanate and ceftriaxone. Microb Drug Resist. 2001;7:85–96.

53. Salih KM, Bilal JA, Eldouch W, et al. Assessment of Treatment of Community Acquired Severe Pneumonia by Two Different Antibiotics. J Clin Diagn Res. 2016;10:SC06–9.

54. Sánchez ME, Gómez J, Gómez Vargas J, et al. Prospective and comparative study between cefuroxime, ceftriaxone and amoxicillin-clavulanic acid in the treatment of community-acquired pneumonia. Rev Esp Quimioter. 1998;11:132–8.

55. Wubbel L, Muniz L, Ahmed A, et al. Etiology and treatment of community-acquired pneumonia in ambulatory children. Pediatr Infect Dis J. 1999;18:98–104.

56. Abid Yaqub ZUK. Comparison of early intravenous to oral switch amoxicillin/clavulanate with parenteral ceftriaxone in treatment of hospitalized patients with community acquired pneumonia. Pak J Med Sci Q. July-September 2005;21:259–66.

57. Rhedin S, Kvist B, Caffrey Osvald E, et al. Penicillin V versus amoxicillin for pneumonia in children-a Swedish nationwide emulated target trial. Clin Microbiol Infect. Published Online First: 16 June 2024. doi: 10.1016/j.cmi.2024.06.008

58. Wei J, Uppal A, Nganjimi C, et al. No evidence of difference in mortality with amoxicillin versus co-amoxiclav for hospital treatment of community-acquired pneumonia. J Infect. 2024;88:106161.

59. Postma DF, van Werkhoven CH, van Elden LJR, et al. Antibiotic treatment strategies for community-acquired pneumonia in adults. N Engl J Med. 2015;372:1312–23.

60. Sulis G, Sayood S, Katukoori S, et al. Exposure to World Health Organization’s AWaRe antibiotics and isolation of multidrug resistant bacteria: a systematic review and meta-analysis. Clin Microbiol Infect. 2022;28:1193–202.

61. Barratt S, Bielicki JA, Dunn D, et al. Amoxicillin duration and dose for community-acquired pneumonia in children: the CAP-IT factorial non-inferiority RCT. Health Technol Assess. 2021;25:1–72.

62. Ginsburg Amy-Sarah, Mvalo Tisungane, Nkwopara Evangelyn, et al. Amoxicillin for 3 or 5 Days for Chest-Indrawing Pneumonia in Malawian Children. N Engl J Med. 2020;383:13– 23.

63. McCallum GB, Fong SM, Grimwood K, et al. Extended Versus Standard Antibiotic Course Duration in Children <5 Years of Age Hospitalized With Community-acquired Pneumonia in High-risk Settings: Four-week Outcomes of a Multicenter, Double-blind, Parallel, Superiority Randomized Controlled Trial. Pediatr Infect Dis J. 2022;41:549–55.

64. Pernica JM, Harman S, Kam AJ, et al. Short-Course Antimicrobial Therapy for Pediatric Community-Acquired Pneumonia: The SAFER Randomized Clinical Trial. JAMA Pediatr. 2021;175:475–82.

