## Supplementary Information for "Effectiveness of amoxicillin and amoxicillin-clavulanate for the treatment of community-acquired pneumonia in adults and children: systematic review and meta-analysis"

**Supplementary Table 1. A.** Characteristics of the 27 included studies with an amoxicillin arm. **B.** Characteristics of the 17 included studies with an amoxicillin-clavulanate arm.

| **Author, Publication Year, Country** | **Study Design** | **Setting** | **Patients** | **Sample Size** | **Daily dose, Route, and Duration of Amoxicillin** | **Daily dose, Route, and Duration of Comparator(s)** | **Extracted Outcomes** |
|---|---|---|---|---|---|---|---|
| CATCHUP Study Group, 2002, Pakistan [13] | randomized controlled, double blind, multicenter | Outpatient | Children <5 with non-severe CAP | 1471 | Oral amoxicillin: 50 mg/kg, 7d | Oral co-trimoxazole: 8/40mg/kg, 7d | Clinical resolution |
| Addo-Yobo, 2004, 8 countries in Africa/Asia/ S. America [14] | Multicenter, randomized, open-label equivalency | Inpatient | Children <5 with severe CAP | 1702 | Oral amoxicillin: 45 mg/kg, 48h min. | Injectable penicillin: 200000 IU/kg, 48h min. | Clinical failure |
| Agweyu, 2015, Kenya [15] | Open-label, multicenter, randomized controlled noninferiority | Inpatient | Children <5 with severe CAP | 521 | Oral amoxicillin: 80-90 mg/kg, 48h minimum | i.v./i.m. benzyl penicillin: 200000 IU/kg, 48h min. | Clinical failure |

| | | | | | | | |
|---|---|---|---|---|---|---|---|
| Ahmed, 2022, Pakistan [16] | Cluster-randomized, unblinded, community-based | Outpatient | Children <5 with fast-breathing CAP | 4984 | Oral amoxicillin: 250mg/5ml, 3d | Oral co-trimoxazole: 200mg/40mg per 5ml, 5d | Clinical failure |
| Aubier, 1998, 46 centres in France/ South Africa/ Switzerland [17] | randomized, double-blind, multicenter | Inpatient | Adults hospitalised with CAP | 329 | Oral amoxicillin: 3000g, 10d | Oral sparfloxacin: 400mg loading, 200mg, 10d | Clinical resolution |
| Awasthi (PLoS), 2008, India [18] | Multi-centric, randomized placebo controlled double blind clinical | Outpatient | Children <5 with non-severe CAP | 1671 | Oral amoxicillin: 31-54mg/kg, 3d | Placebo: 3d | Clinical resolution |
| Awasthi (JTP), 2008, India [19] | Cluster randomized, open labelled | Outpatient | Children <5 with non-severe CAP | 2009 | Oral amoxicillin: 31-51 mg/kg, 3d | Oral co-trimoxazole: 7-11mg/kg, 5d | Clinical failure |
| Baumgartner, 1984, Switzerland [20] | randomized, controlled | Inpatient | Adults with acute CAP | 1984 | i.v. amoxicillin: 4g, duration varied | i.v. ceftriaxone, 2g, duration varied | Clinical resolution |
| Carbon, 1992, France [21] | Double-blind, multicenter | Inpatient | Adults hospitalised with CAP | 246 | Oral amoxicillin: 1000mg, 10d mean | Oral temafloxacin: 1200mg, 10d mean | Clinical failure |
| Ginsburg, 2019, Malawi [22] | Double-blind, randomized clinical noninferiority | Outpatient | Children <5 with non-severe fast-breathing CAP | 1126 | Oral amoxicillin: 500-1500mg, 3d | Placebo: 3d | Clinical failure |
| Hagberg, 2002, 13 countries in S. America/Oceania/ Europe/Africa [23] | randomized, double-blind | Mixed | Adults with CAP | 404 | Oral amoxicillin: 3000mg, 10d | Oral telithromycin: 800mg, 10d | Clinical resolution |
| Hazir, 2008, Pakistan [24] | randomized, open-label, equivalency | Mixed | Children <5 with severe CAP | 2037 | Oral amoxicillin: 80-90 mg/kg, 5d | Parenteral ampicillin: 100mg/kg, 3d | Clinical failure |

| | | | | | | | |
|---|---|---|---|---|---|---|---|
| Hazir, 2011, Pakistan [25] | Double-blind, randomized, equivalence | Inpatient | Children <5 with non-severe CAP | 873 | Oral amoxicillin: 45 mg/kg, 3d | Placebo: 3d | Clinical failure |
| Jardim, 2003, Latin America [26] | Prospective, multicenter, multinational, controlled, randomized, double-blind | Mixed | Adults with CAP | 70 | Oral amoxicillin: 1500mg, 10d maximum | Oral moxifloxacin: 400mg, 10d | Clinical resolution, microbiologic cure |
| Jehan, 2020, Pakistan [27] | Double-blind, randomized, placebo-controlled noninferiority | Outpatient | Children <5 with non-severe CAP | 4002 | Oral amoxicillin: 1000-3000mg, 3d | Placebo: 3d | Clinical failure |
| Leuenberger, 1983, Switzerland [28] | randomized double blind controlled | Inpatient | Adults with CAP | 34 | Oral amoxicillin: 1500mg, 7d | Oral cefaclor: 2250mg, 7d | Clinical resolution |
| Llor, 2019, Spain [29] | Multicenter, parallel, double-blind, controlled, randomized | Outpatient | Adults with CAP | 43 | Oral amoxicillin: 3000mg, 10d | Oral penicillin: 1.6 million units, 10d | Clinical resolution |
| Müller, 1992, Canada/Europe/South Africa [30] | Double-blind, randomized, parallel treatment | Outpatient | Adults with CAP | 336 | Oral amoxicillin: 1500mg, 10-14d | Oral loracarbef: 800mg, 10-14d | Clinical resolution |
| O'Doherty, 1997, UK/Ireland [31] | randomized, multicenter, double-blind, double-dummy | Outpatient | Adults with CAP | 264 | Oral amoxicillin: 1500mg, 7-10d | Oral grepafloxacin: 600mg, 7-10d | Clinical resolution |
| Petitpretz, 2001, 20 countries in S. America/Europe/ Asia/N. America/ South Africa [32] | Multinational, multicenter, double-blind, randomized | Mixed | Adults with CAP | 408 | Oral amoxicillin: 3000mg, 10d | Oral moxifloxacin: 400mg, 10d | Clinical resolution |

| | | | | | | | |
|---|---|---|---|---|---|---|---|
| Rahlwes, 1988, Berlin [33] | Prospective, randomized, comparative | Inpatient | Adults with CAP | 33 | Oral amoxicillin: 2250mg, 9.4d mean | Roxithromycin: 300mg, 9.3d mean | Clinical resolution |
| Rajesh, 2013, India [34] | randomized, controlled | Outpatient | Children <5 with non-severe CAP | 204 | Oral amoxicillin: 40mg/kg, 5d | Co-trimoxazole: 8mg/kg trimethoprim, 5d | Clinical resolution |
| Sadruddin, 2019, Pakistan [35] | Unblinded, cluster-randomized, controlled-equivalency | Outpatient | Children <5 with fast-breathing CAP | 15662 | Oral amoxicillin: 50mg/kg, 3d | Oral co-trimoxazole: 8/40mg/kg, 5d | Clinical failure |
| Straus, 1998, Pakistan [36] | randomized, controlled | Inpatient | Children <5 with non-severe or severe CAP | 595 | Oral amoxicillin: 45mg/kg, 3-5d | Co-trimoxazole: 80mg/8mg /kg, 3-5d | Clinical failure |
| Trémolières, 1998, Europe/ South Africa/ Costa Rica [37] | randomized, multicenter, double-blind study | Mixed | Adults with CAP | 312 | Oral amoxicillin: 3000g, 7-10d | Oral trovafloxacin: 200mg, 7-10d | Clinical resolution, microbiologic cure |
| Trémolières, 2005, France/Tunisia [38] | Multinational, randomized, double blind, double dummy, non-inferiority | Inpatient | Adults with CAP | 371 | Oral amoxicillin: 3000g, 7-10d | Oral pristinamycin, 3000mg, 7-10d | Clinical resolution |
| Tsarouhas, 1998, US [39] | Prospective, randomized, evaluator-blinded | Outpatient | Children (6m-18y) with CAP | 170 | Oral amoxicillin: 50mg/kg, 2d | i.m. procaine penicillin G: 50,000 units/kg | Clinical failure |

**Supplementary Table 1. B.** Characteristics of the 17 included studies with an amoxicillin-clavulanate arm.

| | | | | | | | |
|---|---|---|---|---|---|---|---|
| Bonvehi, 2003, 45 sites in Argentina, Italy, Mexico, South Africa, Spain, Turkey [40] | Prospective, randomized, investigator-blinded, multicenter | Outpatient | Patients ≥12 with CAP | 327 | Oral amoxicillin-clavulanate: 1550/250mg, 7d | Oral clarithromycin, 1000mg, 7d | Clinical resolution, microbiologic cure |
| Carbon, 1999, 50 centres in Argentina, Europe, South Africa [41] | Double-blind, randomized, three-arm parallel, multicenter | Mixed | Adults with mild-to-moderate CAP | 516 | Oral amoxicillin-clavulanate: 1875mg, 7-10d | Oral levofloxacin: 500mg or 1000mg, 7-10d | Clinical resolution, microbiologic cure |
| Finch, 2002, 10 countries in Europe/Africa/ Asia [42] | Multinational, multicenter with a randomized, open, parallel group design | Inpatient | Adults with CAP | 538 | i.v./oral amoxicillin-clavulanate: oral dose of 1875g, 7-14d | i.v./oral moxifloxacin: oral dose of 400mg, 7-14d | Clinical resolution, microbiologic cure |
| Fogarty, 2002, US [43] | Multicenter, prospective, randomized, investigator-blinded, parallel-group | Inpatient | Patients ≥12 with CAP | 802 | Oral amoxicillin-clavulanate: 1750/250mg, 14d | Oral cefditoren: 400mg or 800mg, 14d | Clinical resolution, microbiologic cure |
| Genné, 1997, Switzerland [44] | Open, prospective, randomized, controlled | Inpatient | Adults with CAP | 112 | i.v./oral amoxicillin-clavulanate: oral dose of 1875g, 10d min. | i.v./oral clarithromycin lactobionate: oral dose of 1000mg, 10d min. | Clinical resolution |
| Higuera, 1996, US/Latin America [45] | Multicenter, investigator-blinded | Outpatient | Patients ≥12 with CAP | 168 | Oral amoxicillin-clavulanate: 1500/375mg, 10d median | Oral cefuroxime axetil: 1000mg, 10d mean | Clinical resolution, microbiologic cure |
| Léophonte, 2004, 102 centres in France/Poland/ South Africa [46] | randomized, multicenter, double-blind, double-dummy, parallel group Phase III | Mixed | Adults with CAP | 254 | Oral amoxicillin-clavulanate: 3000/375mg, 10d | Oral gemifloxacin: 320mg, 7d | Clinical resolution, microbiologic cure |
| Lode, 1995, 9 countries in Europe/Israel [47] | Double-blind, randomized, parallel group | Mixed | Adults with CAP | 808 | Oral amoxicillin-clavulanate: 1500/375mg, 9-10d mean | Oral sparfloxacin: 400mg loading dose, 200mg, 9-10d mean<br>Oral erythromycin: 2000mg, 9-10d mean | Clinical resolution |

| | | | | | | | |
|---|---|---|---|---|---|---|---|
| Lode, 2004, 16 countries in Europe [48] | Double-blind, double-dummy, multicenter, multinational, parallel-group | Inpatient | Adults with CAP | 462 | Oral amoxicillin-clavulanate: 1500/375mg, 5-10d | Oral gatifloxacin: 400mg, 5-10d | Clinical resolution |
| Mouton, 1991, 12 centres in France [49] | randomized, controlled, multi-center | Inpatient | Adults with CAP requiring hospitalisation | 63 | Oral amoxicillin-clavulanate: 2000mg, 5d min. | Oral ciprofloxacin: 1500mg, 5d min.<br>Oral erythromycin: 3000mg, 5d min. | Clinical resolution |
| Oh, 1996, Singapore [50] | randomized trial | Inpatient | Adults with CAP | 48 | i.v./oral amoxicillin-clavulanate: oral dose of 1500/750mg, 7d min. | i.v./oral cefuroxime: oral dose of 500mg, 7d min. | Clinical resolution |
| Paris, 2008, Italy [51] | randomized, open-label, non-inferiority | Outpatient | Adults with CAP | 267 | Oral amoxicillin-clavulanate: 1750/250mg, 7d | Oral azithromycin: 1000mg, 3d | Clinical resolution |
| Rosón, 2001, Spain [52] | Comparative, prospective, randomized trial | Inpatient | Adults with CAP | 378 | i.v./oral amoxicillin-clavulanate, oral dose of 3000/375mg, 10.9d mean | i.v./i.m. ceftriaxone: 1000mg, 10.1d mean | Clinical resolution |
| Salih, 2016, Sudan [53] | Prospective case control, randomized sampling | Inpatient | Children <60 months with severe CAP | 83 | Amoxicillin-clavulanate | Penicillin, Ceftriaxone | Clinical resolution |
| Sánchez, 1998 [54] | Prospective, comparative | Inpatient | Patients ≥12 with moderate-to-severe CAP | 409 | i.v./oral amoxicillin-clavulanate: 2625mg oral, 12d total | i.v./oral cefuroxime: 1500g, 12d total<br>i.v./i.m. ceftriaxone: 2g i.v., 5d i.v. | Clinical resolution |
| Wubbel, 1999, US [55] | Prospective, randomized, unblinded | Outpatient | Children <5 with CAP | 147 | Oral amoxicillin-clavulanate: 40mg/kg, 10d | Azithromycin: 10mg/kg d1, 5mg/kg 4d | Clinical resolution |

| | | | | | | | |
|---|---|---|---|---|---|---|---|
| Yaqub, 2005, Pakistan [56] | Open, randomized, parallel group comparative | Inpatient | Adults with CAP requiring i.v. antibiotics | 50 | i.v./oral amoxicillin-clavulanate: 1875mg oral, 7d | Parenteral ceftriaxone: 1-2g, 7d | Clinical resolution |

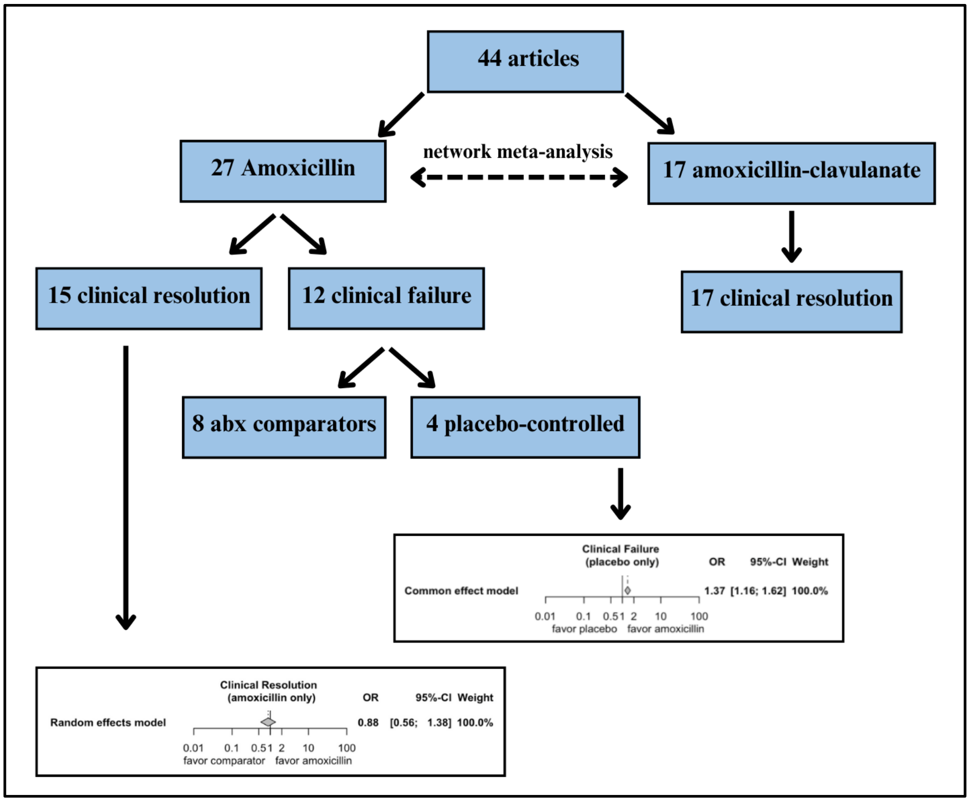


**Supplementary Figure 1.** Flow chart summary of included articles and primary outcomes.

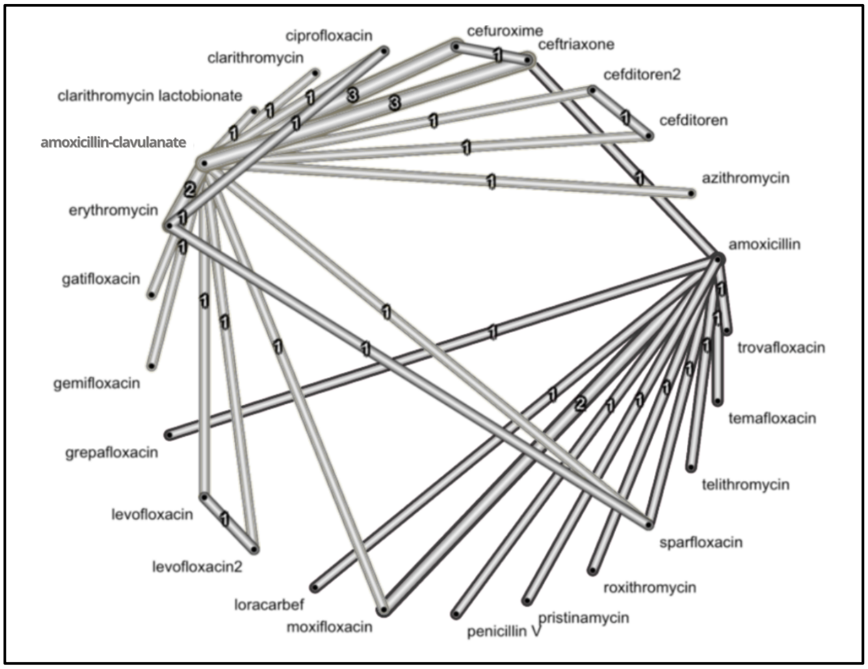


**Supplementary Figure 2.** Network wide meta-analysis of 28 studies considering adult CAP patients with a primary outcome of clinical resolution. Black edges indicate direct comparison between amoxicillin and a comparator antibiotic while grey edges indicate direct comparison between amoxicillin-clavulanate and a comparator antibiotic. The numbers of studies comparing each antibiotic pair are detailed on the edge in the diagram.

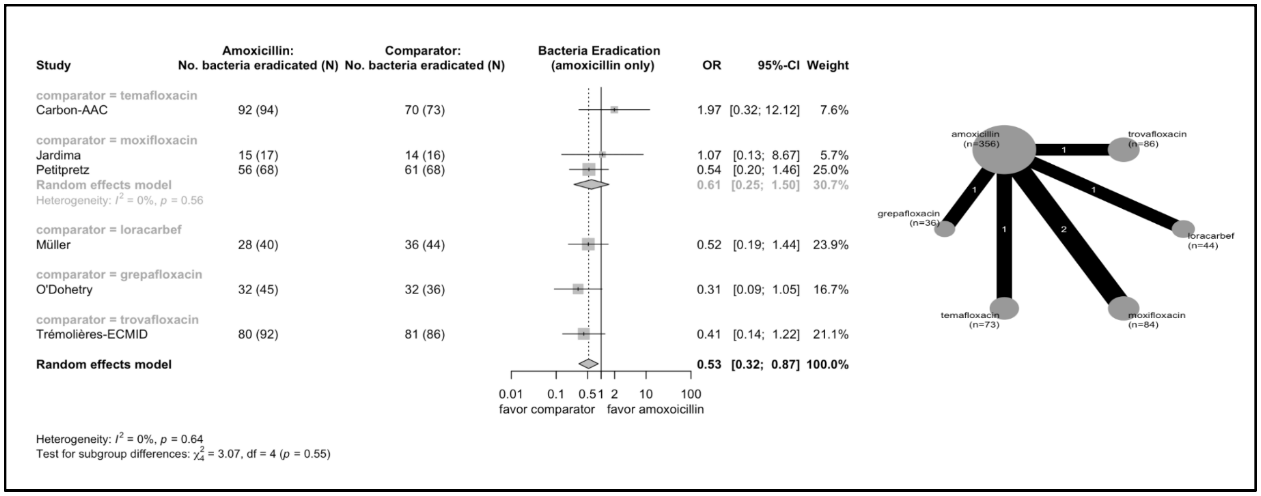


**Supplementary Figure 3.** Forest plot and network graph for six studies with a secondary outcome of presumed or confirmed bacterial eradication comparing amoxicillin to another treatment.

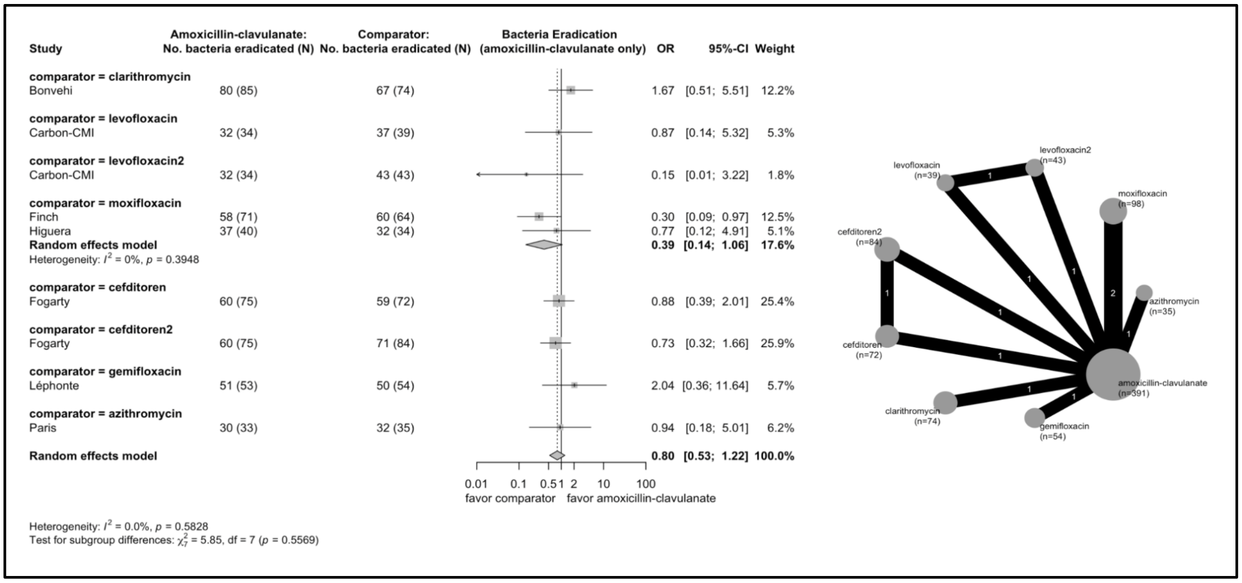


**Supplementary Figure 4.** Forest plot and network graph for seven studies with a secondary outcome of presumed or confirmed bacterial eradication comparing amoxicillin-clavulanate to another treatment.

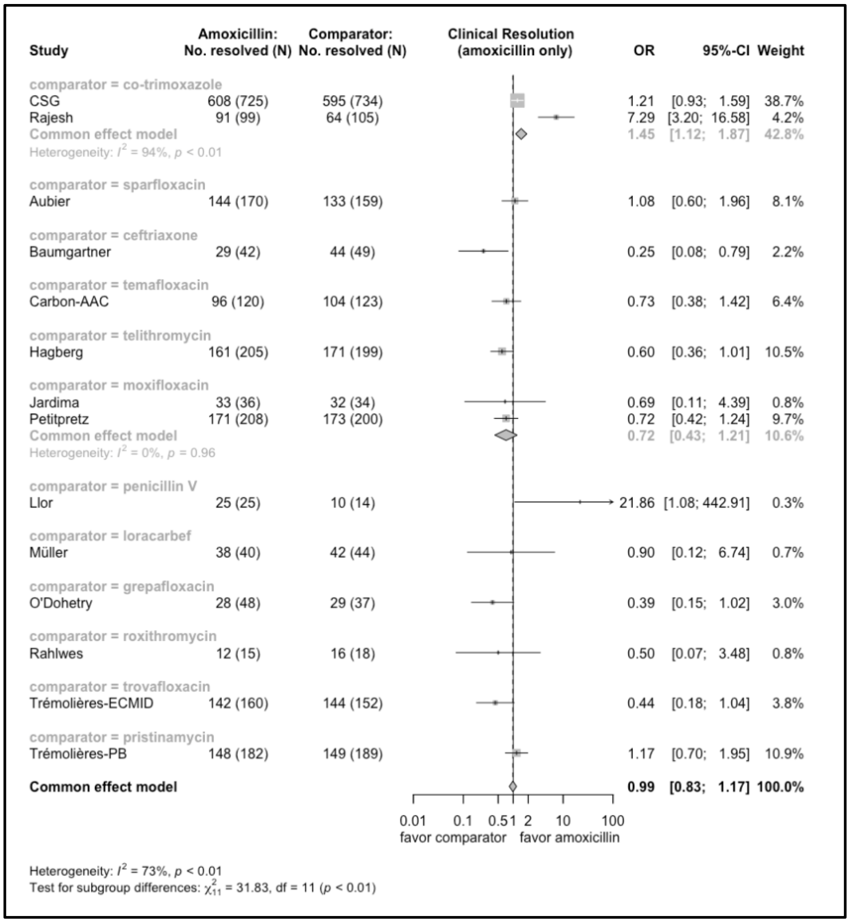

**Supplementary Figure 5.** Forest plot for fixed effects model of clinical resolution, amoxicillin only.

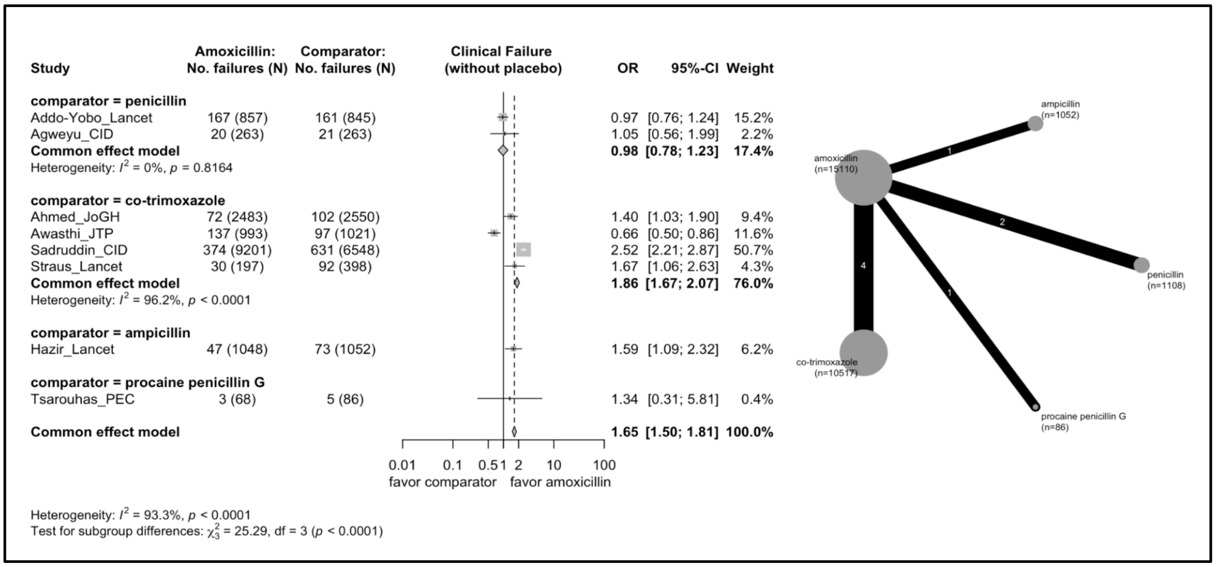


**Supplementary Figure 6.** Forest plot for fixed effects model of clinical failure (without placebo), amoxicillin only. One study (Tsarouhas) considers a patient population of greater than five years of age.

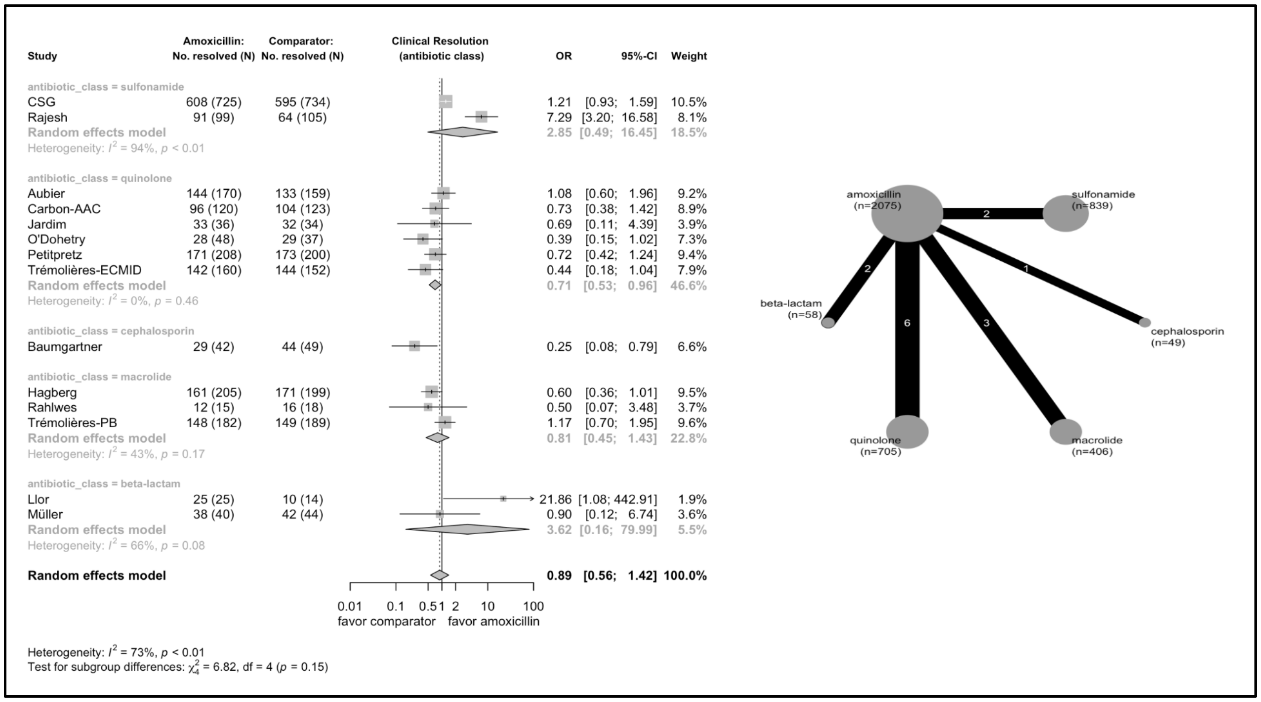


**Supplementary Figure 7**. Forest plot and network graph for clinical resolution by antibiotic class compared to amoxicillin.

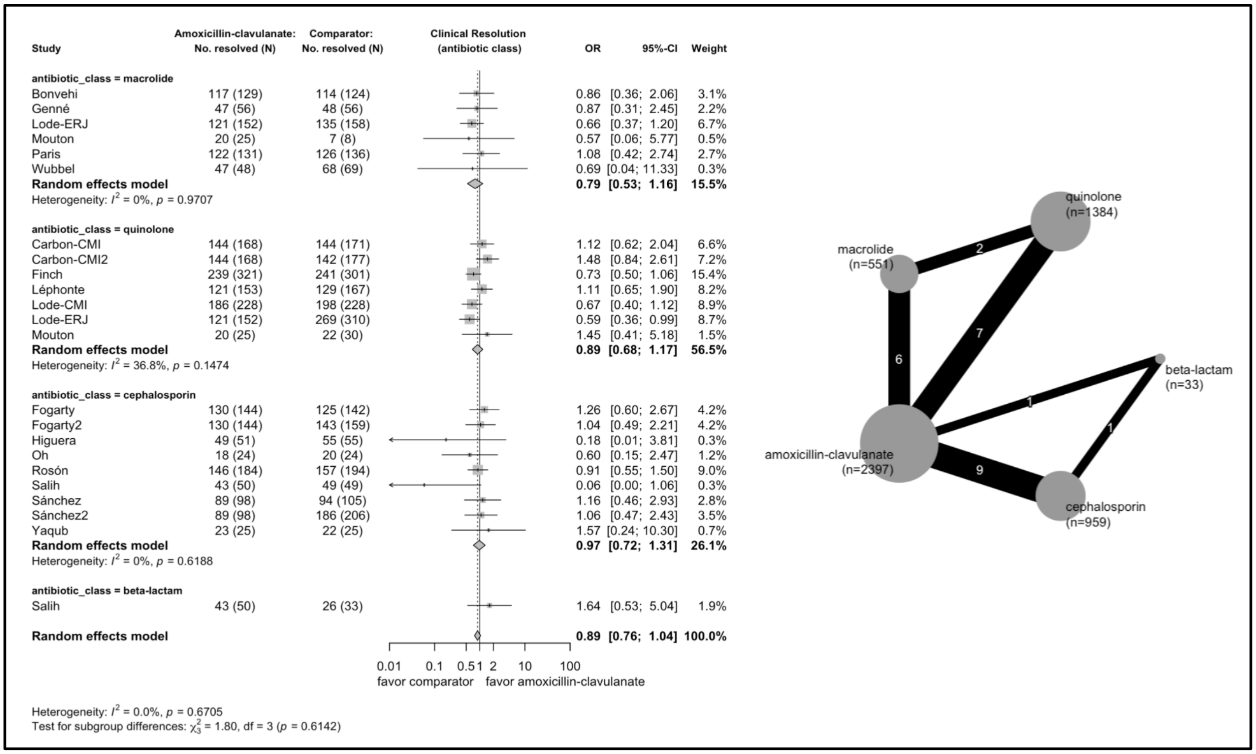


**Supplementary Figure 8**. Forest plot and network graph for clinical resolution by antibiotic class compared to amoxicillin-clavulanate.

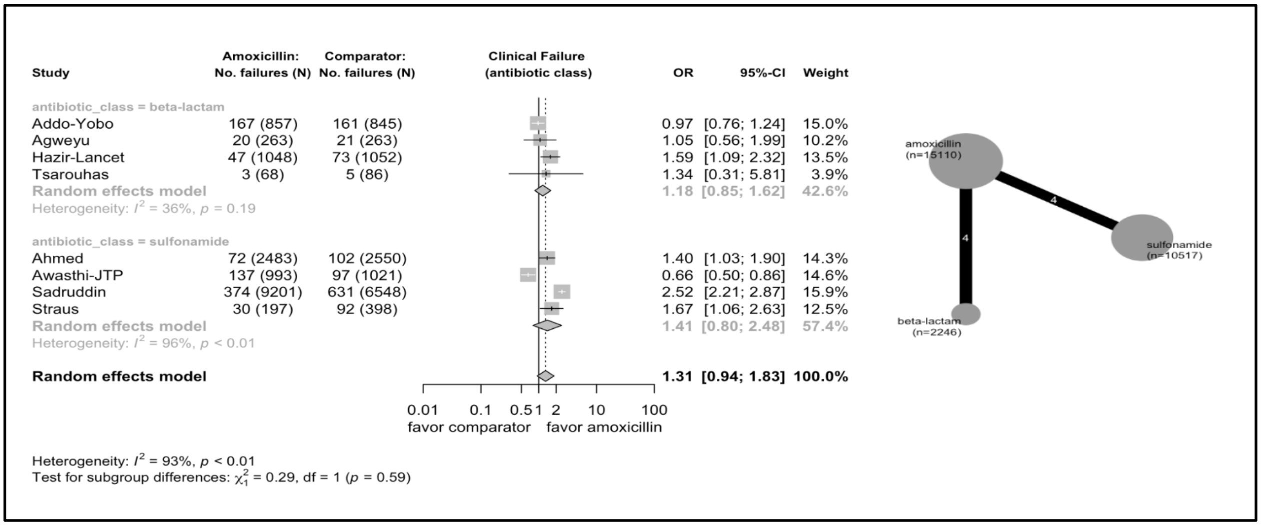


**Supplementary Figure 9**. Forest plot and network graph for clinical failure by antibiotic class (all amoxicillin comparator). One study (Tsarouhas) considered a patient population of greater than five years of age.

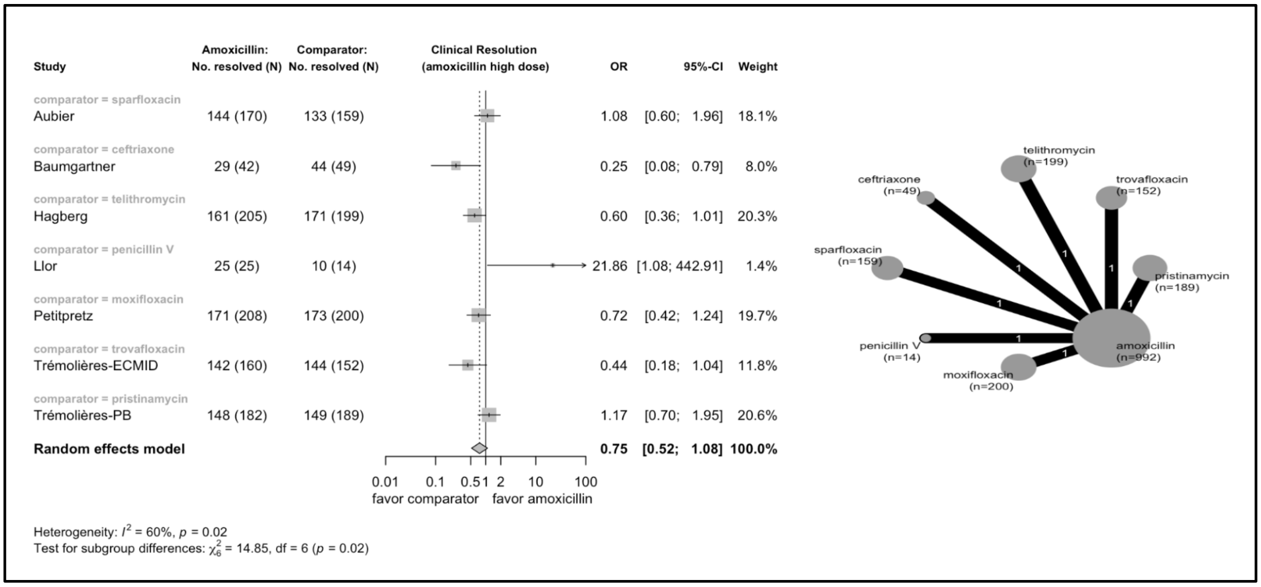


**Supplementary Figure 10.** Forest plot and network graph for clinical resolution and high dose of amoxicillin (≥80 mg/kg/day or 3000 mg/day). All patients were over five years of age.

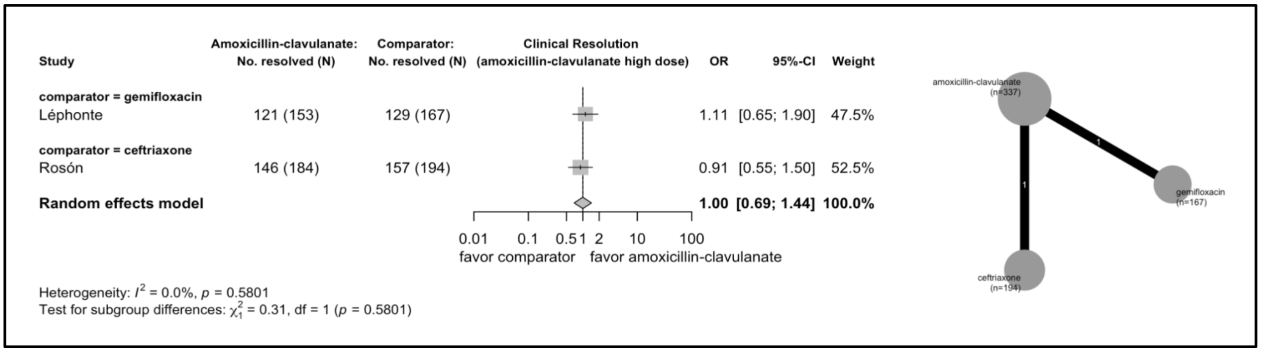


**Supplementary Figure 11.** Forest plot and network graph for clinical resolution and high dose of amoxicillin-clavulanate (≥80 mg/kg/day or 3000 mg/day). All patients were over five years of age.

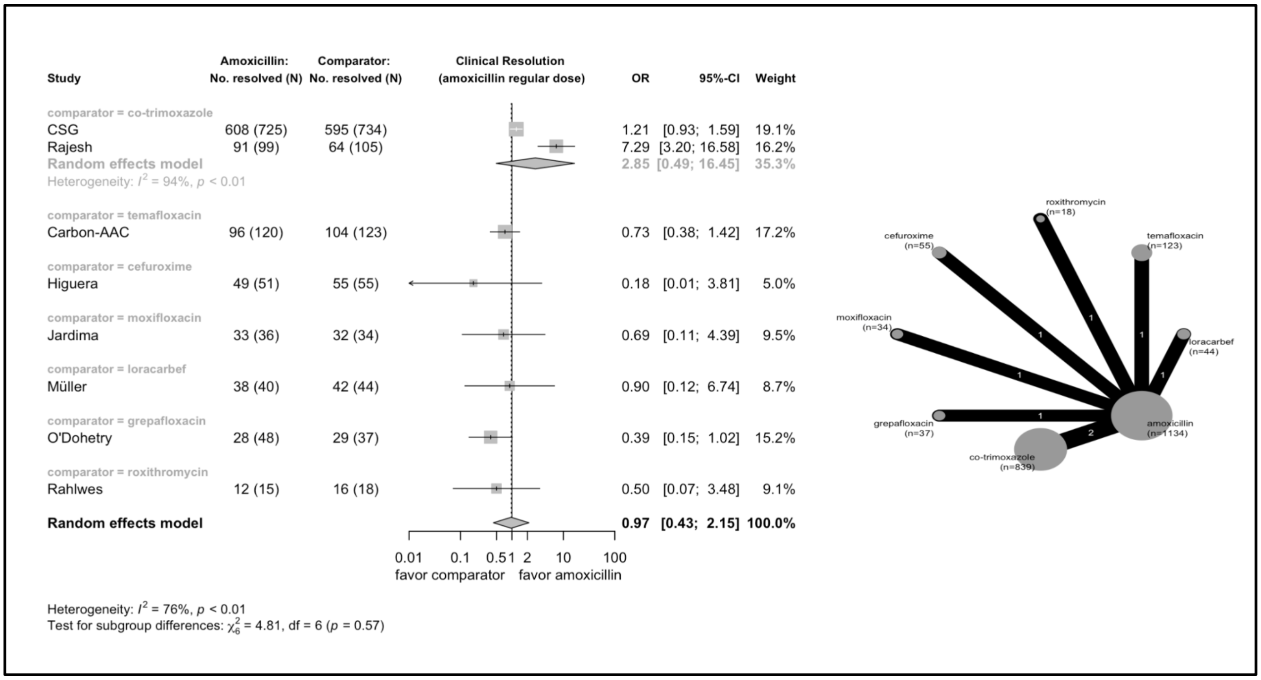


**Supplementary Figure 12.** Forest plot and network graph for clinical resolution and regular dose of amoxicillin. Two studies of patients less than five years of age (CSG, Rajesh).

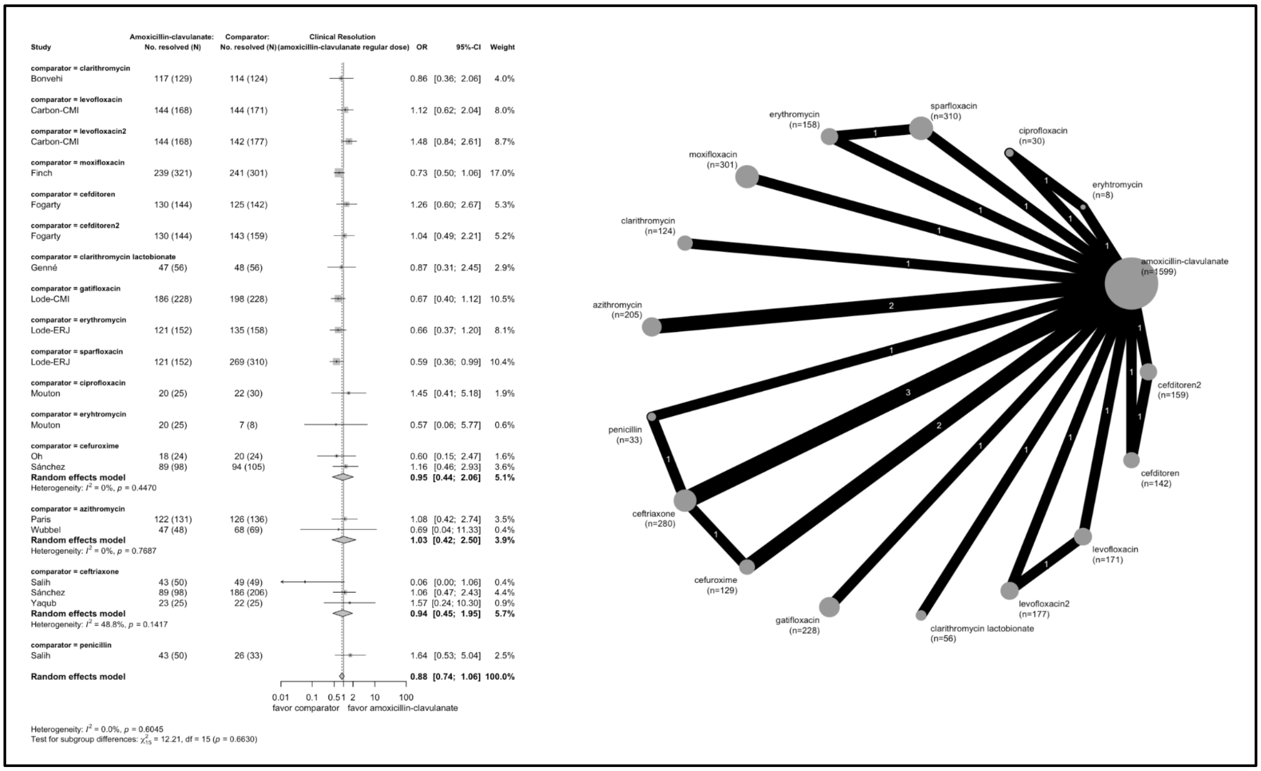


**Supplementary Figure 13.** Forest plot and network graph for clinical resolution and regular dose of amoxicillin-clavulanate. Two studies of patients less than five years of age (Salih, Wubbel).

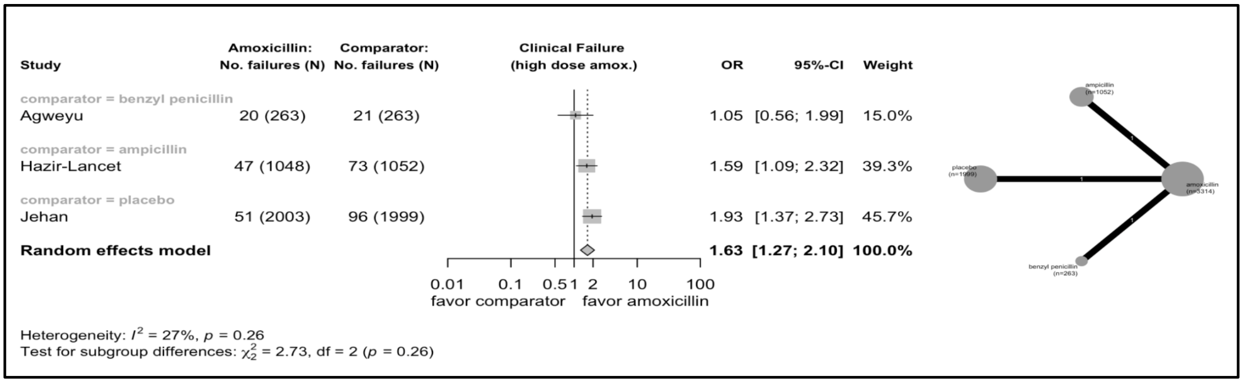


**Supplementary Figure 14.** Forest plot and network graph for clinical failure and high dose of amoxicillin. All studies considered patient populations less than five years of age.

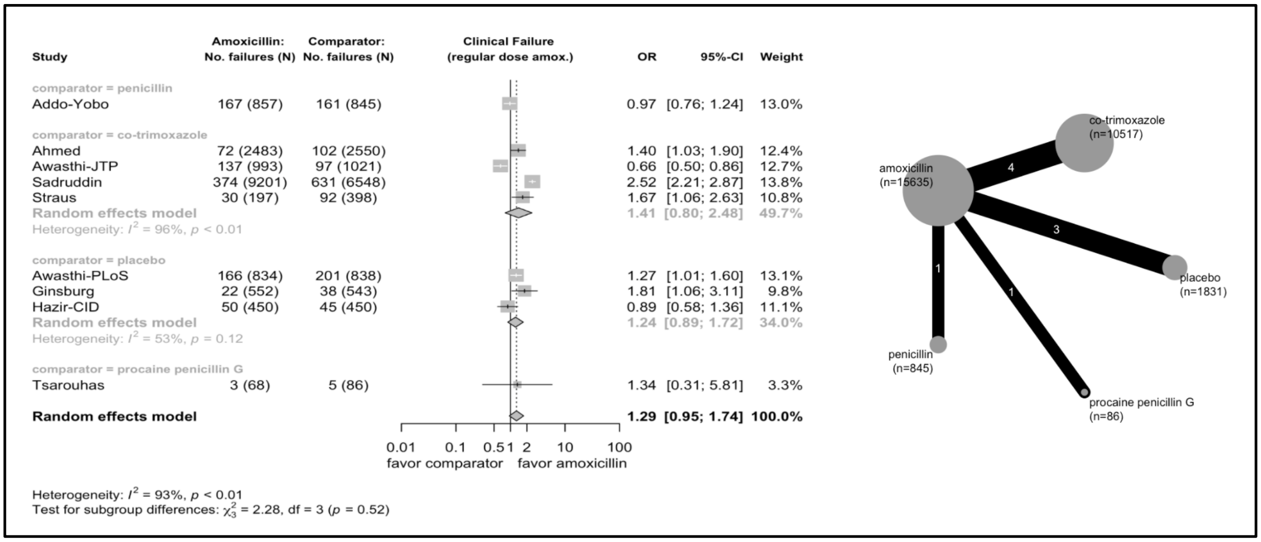


**Supplementary Figure 15.** Forest plot and network graph for clinical failure and regular dose of amoxicillin. One study (Tsarouhas) considered a patient population greater than five years of age.

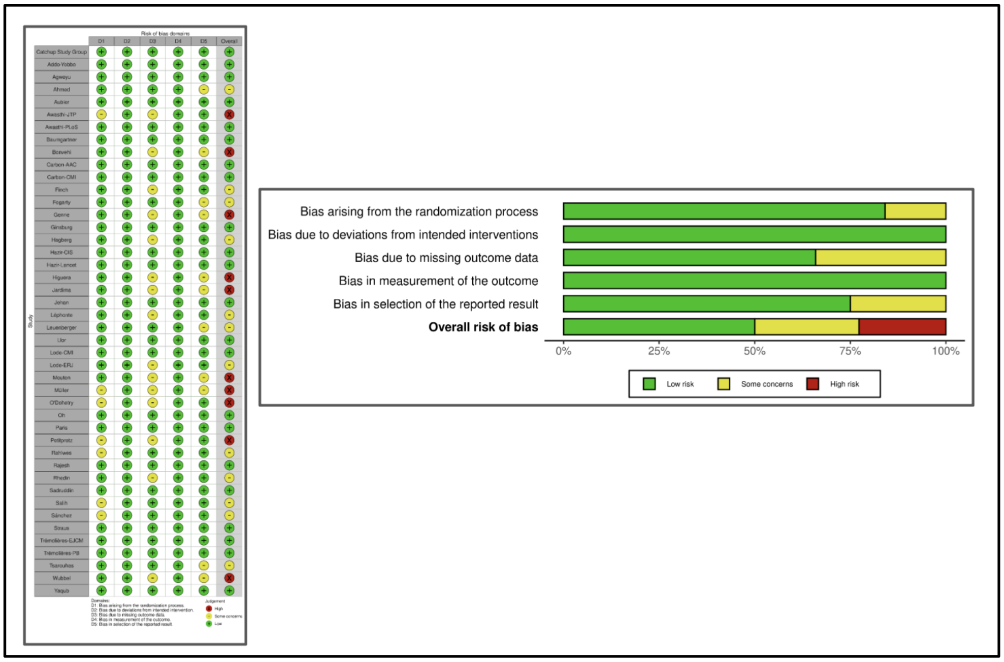


**Supplementary Figure 16.** Risk of bias assessments using the Cochrane RoB 2.0 assessment and R-based shiny web app risk of bias VISualization tool (robvis).
